# Covid-19 impact on food insecurity in Uganda: a dynamic analysis

**DOI:** 10.1101/2023.03.07.23286899

**Authors:** Chisom L. Ubabukoh, Gindo Tampubolon

## Abstract

Lockdowns were used as a tool to avoid excessive social contact and thus limit the spread of Covid-19. However, the true welfare effects of this policy action are still being determined. This paper studies the impact of these lockdowns on the food security outcomes of households in Uganda using a dynamic probit model. We find that the most consequential determinant of whether a household’s food security was severely impacted by the lockdown was the initial status of whether a family was food insecure to begin with. Also, an increase in a household’s economic resources (log consumption per person) significantly influences a reduction in the probability of being severely food insecure. Over time, this creates a wedge of greater inequality between the food security of households who were initially food secure and those who were not. This is despite the use of government cash transfers which have turned out to be ineffective.

**Highlights:**

- A dynamic probit model is used to assess the influence Covid lockdowns have had on food security
- Households who were initially severely food insecure experienced greater levels of food insecurity post-lockdown, than those who were not.
- Increased command of economic resources reduces the probability of severe food security
- Contemporaneous government transfers have not made a significant impact on reducing the probability of severe food insecurity

## 1. Introduction

The tolls of covid-19 on the lives and livelihoods of many households have been devastating around the world, especially on those in the global south as they rely mainly on informal income sources and contain most of the global poor (IMF, 2020). Billions of people were put under one form of lockdown or another at some point in 2020, preventing many of them from securing livelihoods and food for their families. Uganda issued a general lockdown on 20^th^ March 2020 but the consequences of this on food insecurity experienced by families are yet unknown. The custodian of sustainable development goal 2.1 (“zero hunger”), the United Nations’ Food and Agriculture Organisation, has devised a food insecurity experience scale that enables global comparison to be made for such a time as a pandemic.

Even though the worldwide effects of lockdowns are still being studied, there is evidence that they could lead to a worsening of food security (Beckman et al., 2021; Nguyen et al., 2021; Chakrobarty et al., 2020). The channels through which this could happen include increases in food prices (Singh et al., 2020), the suspension of school feeding and other child development programmes (Alvi & Gupta, 2020), supply chain disruptions (Alvi & Gupta, 2020), and a reduction in earning and productive capacity due to the spread of covid-19 and other communicable diseases (Chakrobarty et al., 2020). On the other hand several papers, especially those who have used high frequency (phone) survey data report the exact opposite: that covid-19 lockdowns have not caused a significant decrease in food security, but an actual improvement (Ibukun & Adebayo, 2021; Deaton & Deaton, 2020; Niles et al., 2020; Wolfson & Leung, 2020). The argument they make is that households are surprisingly resilient and have found ways to adapt either by finding alternative food sources, increased borrowing, through philanthropic activity or via other communal networks (Abdullah et al., 2021; Mahmud & Riley, 2020). This notwithstanding, there is likely to have been a reduction in the quality of food consumed, even though food availability was not seriously affected as households switch from vegetables and other rich foods, to less expensive processed carbohydrates (Ambikapathi et al., 2021; Prapkree et.al., 2021; Niles et al., 2021). Also, using an online survey from 422 respondents, Kansiime et al. (2020) found that two-thirds of the respondents experienced some negative income shock due to the pandemic, as well as reduced food security and nutrition quality, measured with the food insecurity experience scale and the frequency of consumption of nutritionally-rich food.

We study the dynamic consequences of covid-19 and its lockdown in Uganda over five bimonthly rounds of panel surveys from May 2020 to February 2021, offering lessons of relevance to global understanding of the unfolding chronic food insecurity in the wake of the pandemic. The base observation was made with the Uganda Household survey 2019/2020, to which high frequency bimonthly surveys were added. We found considerable chronic levels of food insecurity, which is in line with previous studies (Mugume & Muhumuza, 2021; Rwangire & Muriisa, 2021; Kansiime et al., 2020). Also, using the dynamic features of our model, we discover that two-thirds of those in severe food insecurity in one month are likely to be severely insecure two months later and an increased command of economic resources (measured as log consumption per person) reduces the probability of severe food insecurity. Government transfers were initially reaching those in severe food insecurity but contemporaneous government transfers have not made significant impact on reducing the probability of severe food insecurity. The most consequential determinant of which is the initial status of whether households were food insecure to begin with.

The global food insecurity experience scale aptly captures the dynamics of unfolding food insecurity in Uganda, showing the thick wedge between families who were initially severely insecure compared to those initially secure. Those who were initially secure, one year on had a low probability of being severely food insecure (<10%). But the probability of the other group reached 34%, over the same period. In summary, one year on, the measures to tackle food insecurity are more acutely needed to reverse the damage wreaked by the pandemic, in order to achieve zero hunger.

To the best of our knowledge this study is the first to analyse the food security situation of Uganda in particular as it relates to the lockdown using nationally representative data. Mahmud & Riley (2020) use primary data collected from 114 villages in Western Uganda to analyse the economic impacts of the lockdown in that region and find that household incomes fell by 60% and savings were drawn down, borrowing increased and tended to increase their labour supply for farming activities. But food security declined – there was a 50% increase in the likelihood of respondents missing a meal due to the covid-19 lockdown.

The national coverage of the data used in our study is useful because it coalesces strands of regional experiences into a cohesive whole and paints a broad picture of the food security experiences of households nationwide. One practical way this could come into play is in recognising that a lot of people move around to a more comfortable area during a lockdown, probably moving away from temporary residences to family homes. For instance, school children and other students would normally be expected to have moved to their homes or villages during the lockdown and this would make it more difficult to compare the scale of effects before and after the lockdown. We are also able to address nationally relevant policy.

Our study goes beyond an indication of state levels (i.e. snapshot of severe food insecurity) to examine state dependencies and dynamics. The high frequency survey we use is typical; and many such surveys are ongoing in several countries, for example, Himelein & Kastelic (2021) use them to analyse the socio-economic impacts of Ebola in Liberia, Jones & Balon (2020) use them to track changes in resilience after a natural hazard in Eastern Myanmar, and Palacious-Lopez et al (2021)use them to analyse the impact of covid-19 on the labour market in multiple countries. But our use of the data is original. This paper is the first we know that uses a dynamic model to investigate such an important household outcome as food insecurity. This dynamic model is important due to the frequency of visits or rounds, making state or path dependence critical. Ignoring the dynamics or state dependence is likely to make statistical inference biased and spurious. The application of a dynamic modelling approach on an ongoing high frequency survey, as is used here, is suitable to be used in other ongoing high frequency surveys elsewhere.

## 2. Covid-19 Lockdown in Uganda

Due to the prevailing state of panic in many nations around the world after covid-19 was announced to be a pandemic, many of them imposed lockdowns at varying levels of intensity. The lockdown implemented in Uganda was one of the strictest. First as a precautionary move, public gatherings for worship, weddings, rallies, and so on were suspended from the 20^th^ of March, 2020. Then all schools, universities, border crossings, all transportation and non-essential businesses were shut. This was done just as the first case was officially confirmed on the 22^nd^ of March. From the 30^th^, a total nationwide curfew was further imposed from 7pm until 6:30am every day. By the end of March, a total of 44 covid-19 cases had been officially confirmed.

This lockdown had originally been intended to last for 32 days, but it was extended several times until the 5^th^ of May, when a small number of businesses were allowed to open as a start to the gradual lifting of restrictions. The Ugandan government made some food handouts to about 1.4 million struggling households during this lockdown, but this was seen as widely insufficient in helping the food insecure households meet their food needs. A second lockdown was re-imposed to last for 42 days from the 19^th^ of June, 2021; for which the government also promised to make covid-19 relief cash transfers.

Although the economic and welfare effects of these lockdowns on households are still being studied, the early studies show that it has had adverse effects on the people in several ways from both the demand and supply sides (Rwangire & Muriisa, 2021; Mahmud & Riley, 2020; Kansiime et al., 2020).

## 3. Methodology and Data

### 3.1 Data

The data used by this paper is the 5 rounds of the Uganda High-Frequency Phone Survey (UHFPS) on covid-19 from May 2020 to February 2021. The UHFPS was conducted by the Uganda Bureau of Statistics (UBOS), with technical assistance form the World Bank’s Living Standards Measurement Study (LSMS) program and the Poverty and Equity Global Practice, and was distributed and made accessible via the World Bank’s microdata library. The UHFPS was launched in June 2020 for the purpose of tracking the socio-economic impacts of the covid-19 pandemic and was intended to be carried out monthly for period of 12 months, but ended up being a bi-monthly survey. The sample for the survey came from the original sample of households previously interviewed for the Uganda National Panel Survey (UNPS) in the 2019/20 round, who had mobile phones. There were 2,421 households targeted in the original sample, but the different rounds had a little drop-off for random reasons, but always with an above 95% response rate.

### 3.2 Outcome Variable – The Food Insecurity Experience Scale (FIES)

This paper uses the Food Insecurity Experience Scale (FIES) as the means of dividing households into severely food insecure and only moderately food insecure. This measure is chosen because it has become a standard metric used by the Food and Agricultural Organisation (FAO) arm of the United Nations (UN) to indicate worldwide performance levels as it relates to the 2^nd^ Sustainable Development Goal of eliminating hunger and extreme food poverty by 2030. The FIES contains an eight-item question set, for which the respondents answer either a “yes” or “no”. These questions are listed in Table 1.

**Table 1:**
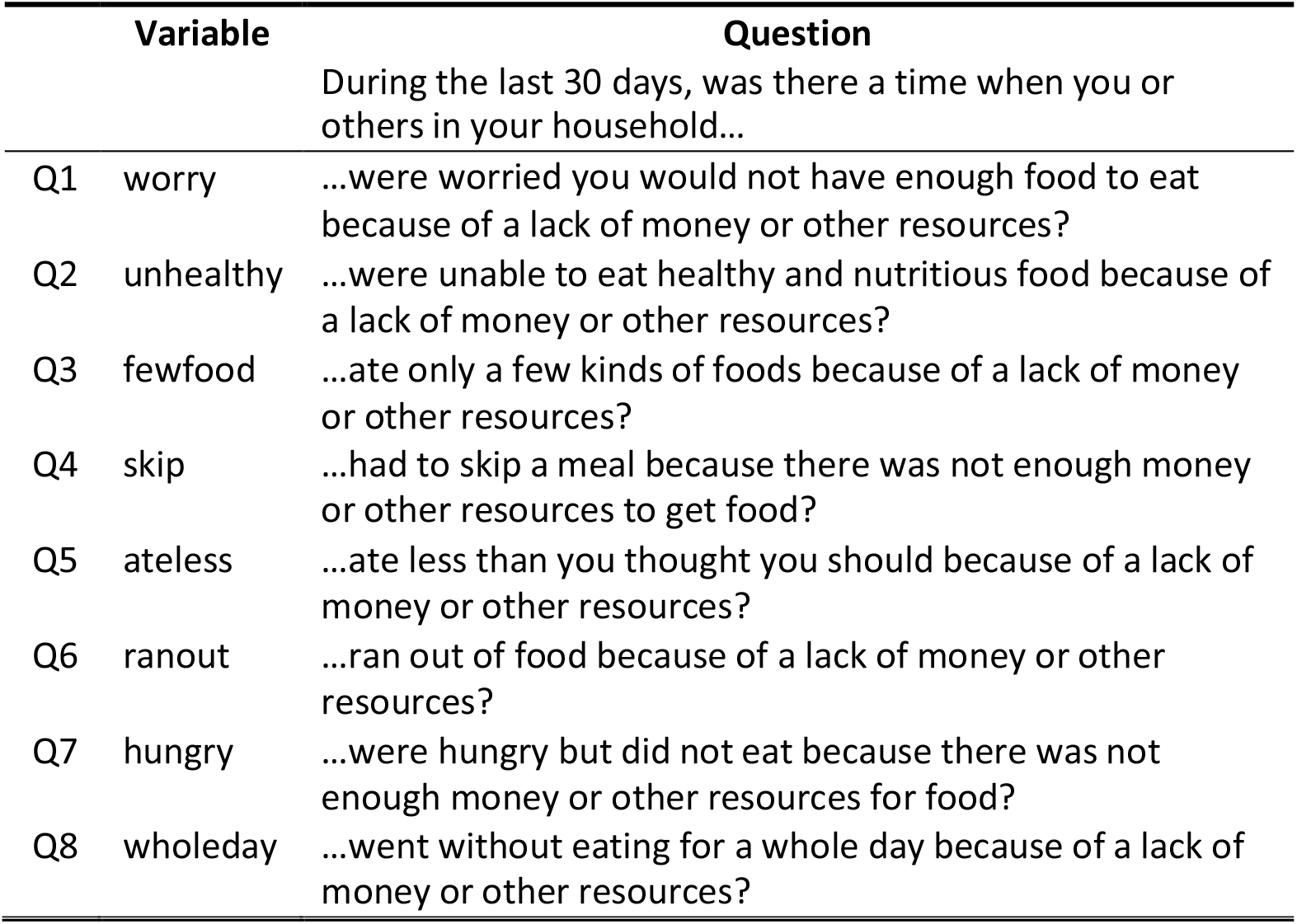
Interview Questions in the Food Security Module of the Ugandan High Frequency Surveys

After the responses are collated, all respondents are given a score from zero to eight. Normally, the FIES score could be recoded as 0 representing food secure, 1-3 representing mild food insecurity, 4-6 for moderate food insecurity and 7-8 for severe food insecurity. However, these simple divisions would be inadequate noting that the context or setting in which the respondent lives could cause great variation in the severity and/or risks of the experience of food security; and thus the raw scores would need to be standardised to some global standard (Cafiero et al., 2018). This is often achieved using Rasch modelling methods (Rasch, 1960) and was done following the technical prescription of the *Voices of the Hungry* project of the FAO (Cafiero et al., 2018). Following the project, users of the scale can derive a binary variable of being in a state of moderate or severe food insecurity versus not, enabling probit modelling of food insecurity status.

### 3.3 Econometric Method

This study uses the dynamic random effects probit model to investigate the main determinants and persistence of severe food insecurity in Ugandan households following the lockdown. This modelling method has been used extensively in the literature to model the determinants and persistence of several economic situations involving discrete choice dynamics including for household welfare and poverty (Alem, 2015), unemployment and underemployment (Hajivassiliou & Ioannides, 2007), and general labour participation (Lee & Tae, 2005; Corsi & Findeis, 2000). Dynamic panel models work well in estimating the impact of a household’s past food securitysituation on the current situation, otherwise known as path dependence. Following Grotti & Cutuli (2018), the model is specified as follows:

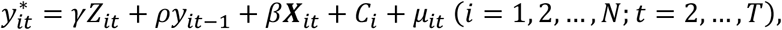

Where 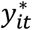 is the latent indicator of severe food insecurity and *y*_*it*_ represents the observed binary outcome variable (severe food insecurity), defined as:

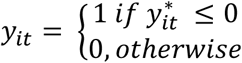

*i* represents households and *t* represents time, *y*_*it*−1_ is the lagged severe food insecurity status of the households used to measure state dependence, ***X***_*it*_ is a vector of explanatory variables, *C*_*i*_ is the unobserved household-specific time-invariant heterogeneity effect, and 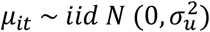 is the error term. The coefficient *γ* represents true state dependence and *β* is a set of associated coefficients to be estimated.

In applying this model here, the incidence of severe food insecurity, which is the outcome variable was regressed on the lagged incidence of severe food insecurity, along with consumption and the other control variables. To estimate the dynamic random effects probit model, there are two main identification challenges that need to be resolved. The first is a problem of initial conditions and the second is that of unobserved heterogeneity. Initial conditions is a problem because of the difference between when the observation is taken in the survey and the start of the development of severe food insecurity which may have been unobserved. The estimates would be biased and inconsistent if there is a correlation between the unobserved condition and the initial observation, but the initial condition was assumed to be exogenous (Heckman, 1981).

The second problem also arises from this fact. There may be some difficulty in differentiating between real statistical path-dependence and the unobserved heterogeneity, especially when they are correlated. There are two main ways that have been suggested to deal with these in the literature: (i) either model the initial response variable together with the ensuing response (Heckman, 1987) or (ii) condition on the initial response by modelling the unobserved heterogeneity relying on the initial dependent variable and other control variables (Wooldridge, 2005). This paper follows the Wooldridge (2005) solution, for which Rabe-Hesketh & Skrondal (2013) show that it could be implemented by including the initial period of the explanatory variables.

Assuming the unobserved heterogeneity is captured by *C*_*i*_, the lagged value of severe food insecurity would represent genuine state dependence. The household-specific unobserved effect can thus be written as:

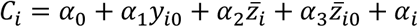

Where *y*_*i*0_ and 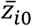 refer to the initial value of the response variable and the time-varying explanatory variables respectively, 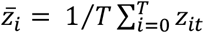 stands for the within-unit averages of the explanatory variables where the averages are based on the available rounds of data; finally, *α*_*i*_ is a household-specific time-constant error term normally distributed with mean 0 and variance 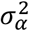.

This strategy was implemented using the “xtpdyn”command in Stata as presented by Grotti & Cutuli (2018).

## 4. Descriptive Statistics

Table 2 shows some descriptive statistics for the 5 rounds on the Uganda high-frequency phone surveys. Of particular interest are the proportions of households in severe food insecurity (which were categorised based on the FIES scores). There was a general improvement in food security outcomes over the survey rounds from about 27% of the households reporting being severely food insecure in the first survey round to only about 8% by rounds 4 and 5, one year on. This is also reflected in figure 1. This is consistent with other research using high frequency (phone) survey data (Hirvonen et al, 2021; Tesfaye et al., 2020; Hirvonen, 2020) and appears to suggest that households have found ways of smoothing food consumption, either by finding alternative food sources, through philanthropic activity, government cash transfer or via other communal networks (Abdullah et al., 2021).

**Table 2:** Summary means of household characteristics for the 5 rounds

| Variables | Round 1 | Round 2 | Round 3 | Round 4 | Round 5 | All |
| --- | --- | --- | --- | --- | --- | --- |
| Severe food insecurity (1 = if household is severely food insecure) | 0.27 | 0.18 | 0.10 | 0.08 | 0.08 | 0.14 |
| Food Insecurity Experience Score (FIES), raw | 3.31 | 2.44 | 1.78 | 1.62 | 1.55 | 2.15 |
| Food Insecurity Experience Score (FIES), adjusted | -0.71 | -1.64 | -2.36 | -2.53 | -2.60 | -1.95 |
| Govt. transfer (1 = if household received a government transfer) | 0.76 | 0.16 | 0.24 | 0.11 | 0.17 | 0.15 |
| Female headed household | 0.33 | 0.33 | 0.33 | 0.33 | 0.33 | 0.33 |
| Household head education (1 = if head has more than secondary education) | 0.34 | 0.34 | 0.34 | 0.34 | 0.34 | 0.34 |
| Age of household head | 48.01 | 48.10 | 48.01 | 48.14 | 48.17 | 48.07 |
| Household size | 4.69 | 4.70 | 4.69 | 4.69 | 4.71 | 4.7 |
| Household consumption | 9811944 | 9729175 | 9680033 | 9790961 | 9737900 | 9750614 |
| Household consumption (per person per day) | 6846.08 | 6757.74 | 6711.46 | 6825.60 | 6731.35 | 6775.25 |
| Household poverty (1 = if household is below poverty threshold) | 0.16 | 0.18 | 0.18 | 0.17 | 0.18 | 0.17 |
| Urban (1 = if household is in an urban area) | 0.26 | 0.26 | 0.26 | 0.26 | 0.26 | 0.26 |
| Number of Observations | 2,217 | 2,147 | 2,094 | 2,086 | 2,069 | 10,613 |

Source: Authors' calculation from the data

**Figure 1:**
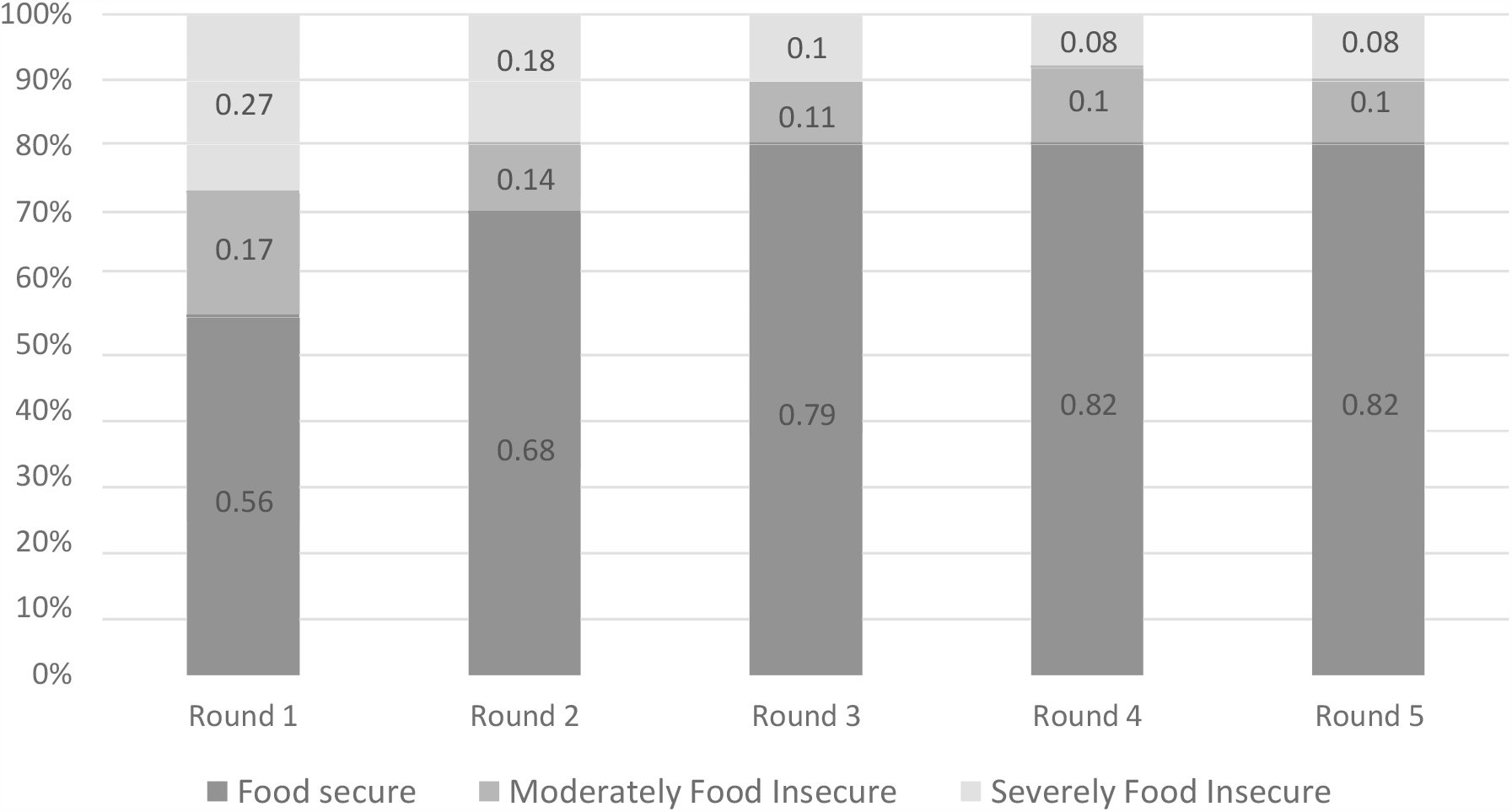
Percentages of food secure, moderately and severely food insecure in the 5 rounds of surveys Source: Authors’ calculation from the data

However, even though there may have been general improvement, one of the main conclusions this paper arrives at by studying the dynamics is that there are significant differences in how this change evolves, depending especially on the initial conditions of the household from before the implementation of the covid-19 lockdown.

On average, the value of household consumption per capita per day in the sample was about 6,775 Ugandan Shillings. This is higher than both the international poverty line of 2,510.4 shillings and the national lower middle-income poverty line of 4,228 shillings, but lower than upper middle-income poverty line of 7,266.9 shillings (Ssewanyana & Okidi, 2007; Duclos et al., 2006), meaning that only about 17% of the households had consumption falling below the international poverty threshold of 1.99 USD per day. One third of the households were female headed, 34% of household heads had above secondary education, 26% lived in urban areas and averagehousehold size was about 5.

From the transitional probabilities reported in table 3, it can be seen that on average, after each round, about 95% of the households that were food secure remained food secure, while only about 5% became severely food insecure. Despite this, the households who were initially severely food insecure had a much lower 57.8% chance of having (or returning to) food security after each round. Also, as reported in table 4, we found that food insecure households who lived in rural areas fared worse with about a 44% chance of remaining food insecure as opposed to the 36% chance for households resident in urban areas. These differences in transitional probabilities are made clearer when the transitional dynamics are examined in the probit model, with a wedge being created between households that were initially food secure and those that experienced severe food insecurity.

**Table 3:** Transitional probabilities of severe food insecurity

| Severe food insecurity, $t$ | Severe food insecurity, $t+1$ | | Total |
| --- | --- | --- | --- |
|  | 0 | 1 |  |
| 0 | 94.99 | 5.01 | 100 |
| 1 | 57.80 | 42.20 | 100 |

Note: The rows reflect the initial percentage values, and the columns reflect the final percentage values.
Source: Authors' calculation from the data

**Table 4:** Transitional probabilities of severe food insecurity divided by urban and rural residence

| Severe food insecurity, $t$ | Severe food insecurity, $t+1$ | | Total |
| --- | --- | --- | --- |
|  | 0 | 1 |  |
| Rural |  |  |  |
| 0 | 94.61 | 5.39 | 100 |
| 1 | 55.95 | 44.05 | 100 |
| Urban |  |  |  |
| 0 | 96.09 | 3.91 | 100 |
| 1 | 64.14 | 35.86 | 100 |

Note: The rows reflect the initial percentage values, and the columns reflect the final percentage values.
Source: Authors' calculation from the data

## 5. Results

The results of the dynamic random effects probit model of severe food insecurity are presented on table 5, column 3. The lagged value of the dependent variable represents state dependence; and having controlled for initial condition and unobserved heterogeneity, we find strong evidence for state dependencies in severe food insecurity. Households who experienced severe food insecurity in one period significantly raises the probability of them being severely food insecure in the subsequent period. Our results indicate that the chances of being severely food insecure increase by 67.9% if the household was severely food insecure in the previous period.

**Table 5:** Results of Random Effects, Probit random effects and Dynamic panel probit random effects models of severe food insecurity

|  | RE<br>(1) | Probit RE<br>(2) | Dynamic Probit RE<br>(3) |
| --- | --- | --- | --- |
| Lag severe food insecurity |  |  | 0.679***<br>[0.0715] |
| Log consumption | -0.685***<br>(0.064) | -0.334***<br>(0.053) | -0.230***<br>[0.0490] |
| Urban | 0.296**<br>(0.108) | 0.192**<br>(0.087) | -0.147*<br>[0.0830] |
| Family led by a woman | 0.260**<br>(0.092) | 0.122*<br>(0.072) | 0.0405<br>[0.0654] |
| Family head has secondary educ. | -0.373***<br>(0.100) | -0.248**<br>(0.081) | -0.110<br>[0.0753] |
| Family head age | 0.0004<br>(0.0028) | 0.001<br>(0.002) | 0.0014<br>[0.0021] |
| Government transfer |  |  | -0.162<br>[0.131] |
| Any work last week |  |  | 0.0023<br>[0.0946] |
| Initially severe food insecurity |  |  | 1.009***<br>[0.0903] |
| Initial government transfer |  |  | 0.422***<br>[0.129] |
| Initial any work |  |  | 0.147*<br>[0.0831] |
| Mean government transfer |  |  | -0.429<br>[0.361] |
| Mean any work |  |  | -0.701***<br>[0.192] |
| *N* | 10,613 | 10,613 | 8,248 |

Note: Standard errors in brackets, constants were included; \* < 0.1; \*\* < 0.05; \*\*\* < 0.001.

Examining the coefficients of the control variables, consumption and living in an urban area were the only other significant determinants of the risk of severe food insecurity. Households who had greater levels of overall consumption had a reduced chance of being food insecure. This result is important because it illustrates the significance having a greater command of economic resources or being non-poor to the food security outcome of the household. This relationship between poverty and food security has been explored in the past (Mahadevan & Hoang, 2016; Zezza & Tasciotti, 2010; Omotesho et al., 2008), but this result provides further evidence of the phenomenon.

Also, we find that urban residents have a *reduced* chance of being food insecure as well. The national scope of the survey data we use allows us to make these useful urban-rural comparisons, and enables a finding like this to be discovered. This is because studies of villagers alone or urban dwellers alone would be precluded from uncovering this information. This result provides further evidence of the urban-rural divide in terms of the effect of shocks on welfare. People in rural areas are less able to cope with shocks such as food insecurity in the wake of covid-19 lockdown. Some papers have suggested (e.g. Chagomoka et al., 2016; Sewnet, 2015) that rural folks can cope by resorting to growing their own food but according to the data here this was wholly inadequate. Direct help would therefore be needed in terms of support for food security outcomes for the rural folks, relative to urban dwellers.

It is worth noting that without dynamic modelling, it appeared that the rural folks were better at coping (urban dwellers have higher probability of being food insecure from the first two models, table 5 columns 1 and 2). But the opposite sign is supported by the transition probabilities above (in tables 4 and 5): people in rural areas are worse off in dynamic terms with worse transition probabilities.

Among the set of variables which capture unobserved heterogeneity or initial conditions, we observe that initial government transfer and initial food insecurity are correlated with unobserved factors positively associated with observed severe food insecurity, in other words, the sample households are also characterised by some time-constant unobserved factors, which increase their risk of severe food insecurity.

Figure 2 shows that households who were food secure initially, had low probabilities of being in severe food insecurity one year on. At any time in the period since the lockdown, their past food insecurity status did not much determine their subsequent food security (flat dark line). In contrast, for those who were initially in severe food insecurity at any time in the period; their initial food insecurity status strongly determined their subsequent food insecurity. Together this difference creates wedge dynamics where the initial difference was amplified in the wake of the pandemic. Government transfers have also failed in preventing the widening gap.

**Figure 2:**
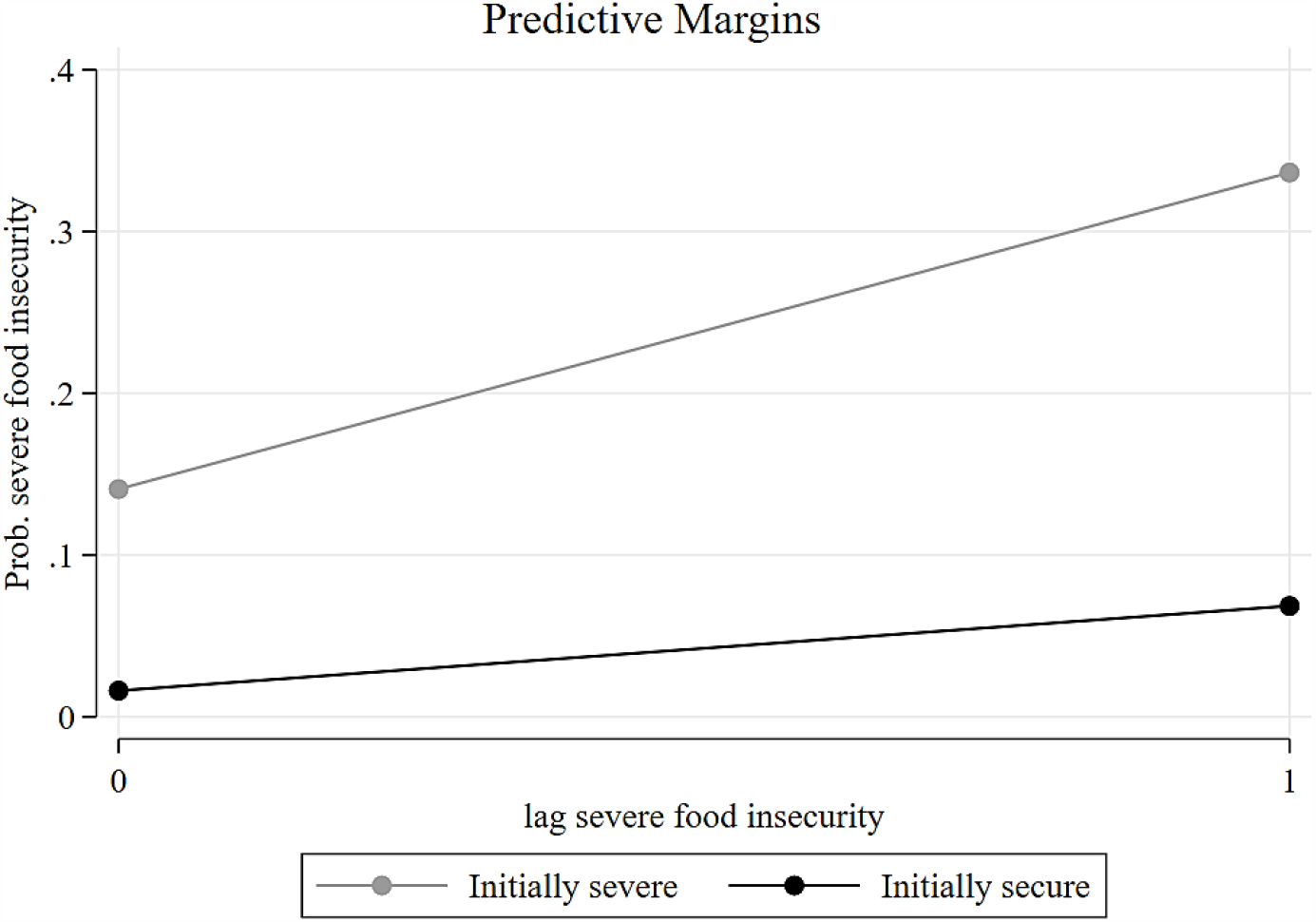
Probabilities of severe food insecurity based on initial food insecurity status and lag of food security (dynamic probit model). Source: Uganda Household Panel Survey 2019/2020 (base) and Uganda High Frequency Surveys rounds 1 to 5 (2020 – 2021)

## 6. Conclusion and Policy Discussion

In conclusion, some governments have been more effective than others in responding to the covid-19 pandemic, devising different degrees of protection to the lives of their people. In Uganda, the food insecurity situation has improved a year on. However, it is equally clear that human development is being hampered by severe food insecurity. If the 2^nd^ SDG of eliminating severe food poverty is to be achieved, more attention needs to be paid not only to the amount of food available as a dimension of food security, but also to the access to food dimension. The most important component responsible for a lack of access to food by households even when it is available (and especially when it is not), is a lack of the economic means to acquire the food. This dimension forces a society to ask itself questions regarding whether an inequality in access to food based on economic resources is appropriate. Our results showed a wedge created between households who initially were food secure and those who were not, and this inequality increased quite dramatically over the survey period.

From the perspective of public health, food equity has been recognised as being both desirable and achievable (Pollard et al., 2016; Baum, 2008) and should therefore be one of the basic objectives of social and economic policy. Some problems associated with chronic hunger including undernutrition, malnutrition and ultimately increased mortality, all have direct impacts on the health outcomes of the population, leading to a loss in human capital and productivity, and eventually to a reduction in the pace of economic growth.

Our use of nationally representative data enables us to draw several general points and policy implications. First, since poorer households and those that have been severely food insecure in the past appear to be more vulnerable to experiencing persistent food insecurity, policy should focus on improving the outcomes of these groups. However, the results also show that government transfers have not so far been an effective way of achieving improved results and there are several reasons why this may be the case. It is possible that estimating the target group was not done properly in the first place and thus resources are wasted or not channelled appropriately. This problem could be solved if the government were more careful in the targeting of transfers to where they would make the greatest improvements. Another reason could be a lack of sustainability; because if households are simply given some money as a one-off transfer and those resources are insufficient to provide long-term food security, the household is then likely to relapse when the funds have run out. To solve this, the government could work out a strategy of systematic transfers in instalments spread out over the relevant period. But even this approach would probably not last forever. From our results, which showed the importance of command of economic resources in achieving long-term food security, in order for vulnerable households to experience food security in a sustainable way, the government would need to adopt appropriate macro-economic policies including proper monetary policy to avoid the negative real income effects triggered by the hyper-inflation of food and other agricultural products, for which the poor spend a disproportionate share of their income (De & Kakar, 2021; Banerjee & Duflo, 2007; Ravallion, 1998). Therefore, the overall economic situation of the households would need to be addressed because, as our results show, food insecurity and poverty appear to be joined at the hip.

## Data Availability

All data used are available online via The World Bank microdata datasets.

https://microdata.worldbank.org/index.php/catalog/3765

